# The influence of highly effective modulator therapies on the sputum proteome in cystic fibrosis

**DOI:** 10.1101/2023.04.17.23288625

**Authors:** Rosemary E Maher, Peter J Barry, Edward Emmott, Andrew M Jones, Lijing Lin, Paul S McNamara, Jaclyn A Smith, Robert W Lord

## Abstract

**Background:** There have been dramatic clinical improvements in cystic fibrosis (CF) patients commenced on the cystic fibrosis conductance regulator (CFTR) modulator elexacaftor/tezacaftor/ivacaftor (ETI). Sputum proteomics is a powerful research technique capable of identifying important airway disease mechanisms. Using this technique, we evaluated how ETI changes the sputum proteome in people with CF.

**Methods:** Sputum samples from 21 CF subjects pre- and post-ETI, 6 CF controls ineligible for ETI, and 15 healthy controls were analysed by liquid chromatography mass spectrometry.

**Results:** Post-ETI, mean FEV_1_% increased by 13.7% (SD 7.9). Principal component and hierarchical clustering analysis revealed that the post-ETI proteome shifted to an intermediate state that was distinct from pre-ETI and healthy controls, even for those achieving normal lung function. Functional analysis showed incomplete resolution of neutrophilic inflammation. The CF control sputum proteome did not alter. At the protein-level many more proteins increased in abundance than decreased following ETI therapy (80 vs 30; adjusted p value <0.05), including many that have anti-inflammatory properties. Of those proteins that reduced in abundance many were pro-inflammatory neutrophil-derived proteins. Several important respiratory proteases were unchanged.

**Conclusions:** Sputum proteomics can provide insights into CF lung disease mechanisms and how they are modified by therapeutic intervention, in this case ETI. This study identifies imbalances in pro- and anti-inflammatory proteins in sputum that partially resolve with ETI even in those achieving normal spirometry values. This post-ETI intermediate state could contribute to ongoing airway damage and therefore its relevance to clinical outcomes needs to be established.

## INTRODUCTION

Cystic fibrosis (CF) is an inherited, multi-system disease caused by dysfunction of the cystic fibrosis transmembrane conductance regulator (CFTR) protein, a ubiquitous ion channel important for epithelial hydration. A direct consequence of this dysfunction is impaired mucociliary clearance, chronic airway infection and a persistent neutrophilic inflammatory response that results in progressive loss of lung function, development of respiratory failure and premature death [1]. Partial restoration of CFTR function is now possible for most CF patients through mutation specific CFTR modulators. Ivacaftor monotherapy produces significant clinical improvement in CF patients with the *G511D* mutation[2]. Dual therapy, combining ivacaftor with lumacaftor or tezacaftor, results in modest clinical improvements in patients homozygous for *F508del* [3, 4]. More recently, triple therapy with elexacaftor/tezacaftor/ivacaftor (ETI) has led to dramatic improvements in lung function and quality of life in patients homozygous and heterozygous for *F508del* [5-7].

Sputum proteomics is a powerful research technique capable of identifying important airway disease mechanisms by interrogating the proteome, an entire set of proteins within biological samples. It has confirmed the central role of neutrophilic immune dysregulation in CF and non-CF bronchiectasis, particularly involving the release of antimicrobial proteins and neutrophil-extracellular traps (NETs), and through impaired anti-inflammatory mechanisms [8-10]. These processes produce distinct molecular signatures within the sputum proteome that become increasingly abnormal with chronic airway infection and progressive lung disease severity [10-13]. In CF patients, airway and systemic inflammatory cytokines potentially related to these signatures reduce with the various forms of CFTR modulation [14-19]. To date, no studies of ETI therapy in CF lung disease have assessed large-scale change in protein expression using untargeted proteomics.

We hypothesised that ETI therapy would shift the sputum proteome toward health, potentially normalising airway biology in people with CF. The objectives of this study were to investigate changes in the CF sputum proteome with the introduction of ETI, correlate these with changes in clinical markers of disease severity, and make comparisons with the sputum proteome in healthy controls and in repeat samples from CF patients not suitable for ETI therapy. We also explored which molecular pathways associated with CF lung disease did not change with ETI. Some of the results of these studies have been previously reported in the form of an abstract [20].

## MATERIALS AND METHODS

### Study design

This prospective observational single-centre study enrolled CF subjects at a large UK specialist CF centre and was conducted under the ethical approval granted to the Manchester Allergy, Respiratory and Thoracic Surgery (ManARTS) Biobank by the Northwest Haydock Research Ethics Committee (20/NW/0302). Sputum from CF subjects was collected pre- and post-ETI for proteomic analysis: post-ETI refers to after ETI was commenced and ongoing at the time of follow-up. These subjects are referred to as the ETI-CF cohort. We combined CF patients who were established on dual CFTR modulators pre-ETI, with those that were modulator therapy naïve. For comparison, we collected longitudinal samples from a CF disease control group (Control-CF) ineligible for modulator therapy based on genotype, and healthy control samples (Supplementary Figure 1). All subjects provided written informed consent. Fully anonymised Study IDs were provided that cannot be identified outside the research group. Subjects were aged over 18 years, had a confirmed diagnosis of CF using genetic testing and/or sweat testing with typical phenotypic features and were deemed clinically stable by the medical team. Exclusion criteria included pregnancy and organ transplantation. The healthy control group comprised adult non-smokers with no history of respiratory disease or evidence of airway obstruction (FEV_1_% predicted ≥ 80% and FEV_1_/FVC ratio ≥ 0.7).

### Sample collection

Spontaneous sputum samples were collected immediately prior to starting ETI and then repeated at the first follow-up clinical visit, typically occurring at 3 months, but up to one-year post-commencing ETI. The interval for CF control group sampling was also 3 -12 months. This window was wide due to COVID restrictions within our clinic. Samples were taken when subjects attended an outpatient assessment and were determined clinically stable by the medical team. Induced sputum samples were collected from healthy control subjects as described previously [10]. All samples were stored at -80°C prior to analysis.

### Proteomic analysis

Sputum samples were processed and underwent tryptic digestion. Samples were analysed by liquid chromatography mass spectrometry (LCMS) using an Ultimate 3000 nano LC system coupled to QExactive-HF mass (Thermo Fisher Scientific, Hemel Hempstead, UK) (see supplementary methods).

### Clinical data

All routine clinical data was collected, including spirometry which was performed either in clinic or at home using the Bluetooth® Air Next Spirometer device (Nuvoair, Stockholm, Sweden) [21].

### Statistical and bioinformatic analysis

Raw LCMS data were processed using Progenesis. Downstream data analysis was performed with a series of bioinformatic approaches (Supplementary figure 2) using R (version 4.0.3). The effects of ETI on the entire proteome relative to healthy controls and CF disease controls were evaluated by: 1) principal component analysis (PCA) using the *prcomp* package; 2) hierarchical clustering using *heatmap3* package. Change with ETI at the individual protein level for CF subjects was tested using paired Mann-Whitney U, with Benjamini-Hochberg correction for multiple comparisons [15]. Functional enrichment analysis of identified proteins was performed using the ShinyGO server [16]. We identified proteins not influenced by ETI therapy using equivalence testing with the two one-sided test (TOST) procedure within the *TOSTER* package, with the equivalence bound specified based on the smallest effect size of interest [22].

## RESULTS

Of the 71 CF subjects enrolled that commenced ETI, 25 provided sputa pre- and post-commencing ETI therapy, whilst 37 were unable to spontaneously expectorate sputum and a further 9 failed to attend their follow up study visit (Supplementary Figure 3). A further four were excluded because of poor in-group sample alignment during data analysis (< 70%) due to low quality LC-MS/MS likely reflecting issues with the original samples (Supplementary Figure 4). Consequently, the study included 42 participants: 21 CF subjects commenced on ETI; 6 CF disease controls (ineligible for ETI based on genotype); and 15 healthy controls. Clinical and demographic characteristics for the cohorts are shown in Table 1, and individual characteristics within Supplementary Table 1. The mean (SD) time from baseline to follow-up sampling was 163 (**±**78) days.

**Table 1.** Demographic and clinical characteristics of patients.

| Demographic | Healthy controls (n=15) | Baseline ETI CF (n=21) | Control CF (n=6) |
| --- | --- | --- | --- |
| Age, mean (SD) | 38.1 (10.7) | 30.9 (7.8) | 36.7 (11.9) |
| Sex, Male (n%) | 3 (20) | 11 (52.4) | 5 (83.3) |
| Baseline FEV<sub>1</sub>%, mean (SD) | 99.7 (9.1) | 63.9 (15.1) | 62.2 (12.7) |
| Body mass index, mean (SD), kg/m<sup>2</sup> | 25.9 (4.9) | 25.3 (5.3) | 18.5 (9.3) |
| Genotype, n (%) |  |  |  |
| Phe508del-Phe508del |  | 13 (61.9) | 0 (0) |
| Phe508del-Minimal function |  | 8 (38.1) | 0 (0) |
| CF-related complications, n (%) |  |  |  |
| Exocrine pancreatic insufficiency |  | 21 (100) | 5 (83.3) |
| CF related diabetes mellitus |  | 7 (33.3) | 1 (16.7) |
| CFTR modulation status, n (%) |  |  |  |
| Tezacaftor/Ivacaftor |  | 10 (47.6) | 0 (0) |
| Lumacaftor/Ivacaftor |  | 2 (9.5) | 0 (0) |
| Long-term medications, n (%) |  |  |  |
| Prednisolone |  | 3 (14.3) | 0 (0) |
| Azithromycin |  | 19 (90.5) | 5 (83.3) |
| Nebulised antibiotic |  | 19 (90.5) | 4 (66.7) |
| Nebulised hypertonic saline |  | 8 (38.1) | 3 (50) |
| Dornase alfa |  | 14 (66.7) | 4 (66.7) |
| Sputum microbiology, n (%) |  |  |  |
| Chronic PsA |  | 20 (95.2) | 4 (66.7) |
| Chronic Bcc |  | 4 (19.0) | 0 (0) |
| Chronic other proteobacteria |  | 3 (14.3) | 0 (0) |
| Chronic NTM |  | 0 (0) | 0 (0) |

Definition of abbreviations: Ps = *Pseudomonas aeruginosa*, Bcc = *Burkholderia cepacia* complex, NTM = *Non-tuberculous mycobacterium*.
Data are presented as absolute number (percentage) or mean (SD)

### Clinical response to ETI

In the ETI-CF group, mean (SD) FEV_1_% increased by 13.7 (7.9)% between starting ETI and the time of repeat sampling (63.3 (15.6)% vs 76.9 (19.2)%, p=<0.001). The percentage of those in the ETI-CF group with normal lung function (FEV_1_≥80%) increased from 9.5% (2/21) to 38.1% (8/21) (p=0.03). In contrast, Control-CF FEV_1_% was unchanged over the follow-up period (62.2 (13.9)% vs 63.0 (13.3)%).

### Identification of core sputum proteome

From the 69 sputum samples (two longitudinal samples from each of the 21 ETI-CF and 6 Control-CF subjects, and one sample from the 15 healthy controls) analysed, 722 proteins were identified based on one or more unique peptide identifications and a false discovery rate of < 1%, i.e. the accepted percentage of expected false protein identifications amongst positive protein identifications. To further increase confidence in protein identification, proteins were further filtered based on a requirement for at least two unique peptides. This resulted in a core sputum proteome of 446 high confidence protein assignments.

### ETI therapy shifts CF toward a healthy sputum proteome

Principal component analysis (PCA) highlighted clear proteome differences, separating the healthy control and pre-ETI sputum samples in the first two principal components without overlap (Figure 1a). The pre-ETI sputum proteomes were very similar for subjects that were modulator naïve (n=9) and taking dual CFTR modulator therapy (n=12), thus justifying combining into a single pre-ETI cohort (Supplementary Figure 5a). Post-ETI, subjects positioned between pre-ETI and healthy control groups consistent with an intermediate state (Figure 1a). When mapping the movement of paired samples onto the PCA, all but one post-ETI sputum sample moved toward the healthy control group (Figure 1b). In contrast, there was movement in both directions in Control-CF subjects with the overall sputum proteome unchanged (Supplementary Figure 5b), suggesting that movement toward health reflects response to ETI therapy. While with progressive lung function impairment the proteome moved away from healthy controls (R = 0.43, p = 0.05; figure 1c), there was no correlation between change in PC1 and change in lung function (R = -0.26, p > 0.05) post-ETI. Timing of the second sample did not influence the sputum proteome (Supplementary Figure 5c), with no significant correlation between time on ETI therapy and change in PC1 (R = 0.24, p > 0.05).

**Figure 1.**
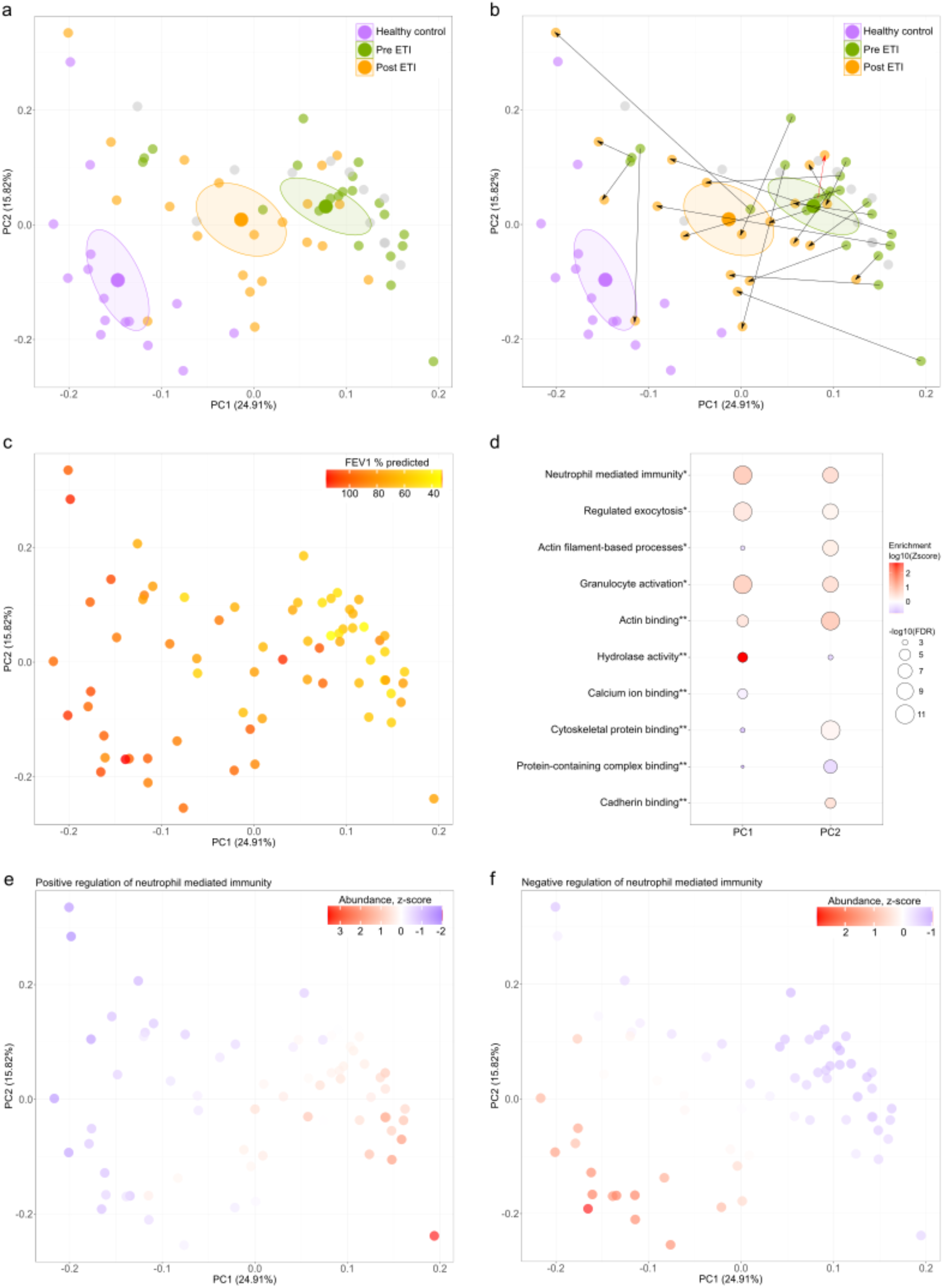
Principal component analysis of sputum protein profiles. (a) Principal component analysis (PCA) of 446 core sputum proteins colour-coded by clinical cohort. A centroid marker (large dot) with 95 % confidence ellipse was calculated for each condition. (b) PCA highlighting movement of paired samples. (c) PCA colour-coded by percent predicted FEV_1_ values. (d) Protein groups identified most enriched *biological process and **molecular functions for PC1 and PC2. PCA plots color-coded by the z-scored abundance of proteins corresponding to (e) positive regulation of neutrophil mediated immunity and (f) negative regulation of neutrophil mediated immunity.

### Proteome differences persist with ETI therapy even in subjects with normal lung function

We performed hierarchical clustering of all participant samples using the top 10% of proteins that were most different between CF and healthy as defined by PC1. CF subjects clustered into two groupings largely independent of modulator status (Figure 2); those with (alpha) and those without (beta) lung function impairment. These clusters remained even when the healthy control cohort was removed (Supplementary Figure 6). Although CF subjects with normal lung function clustered much closer to healthy controls (delta), most remained distinct (gamma) in keeping with persistent differences in the sputum proteome.

**Figure 2.**
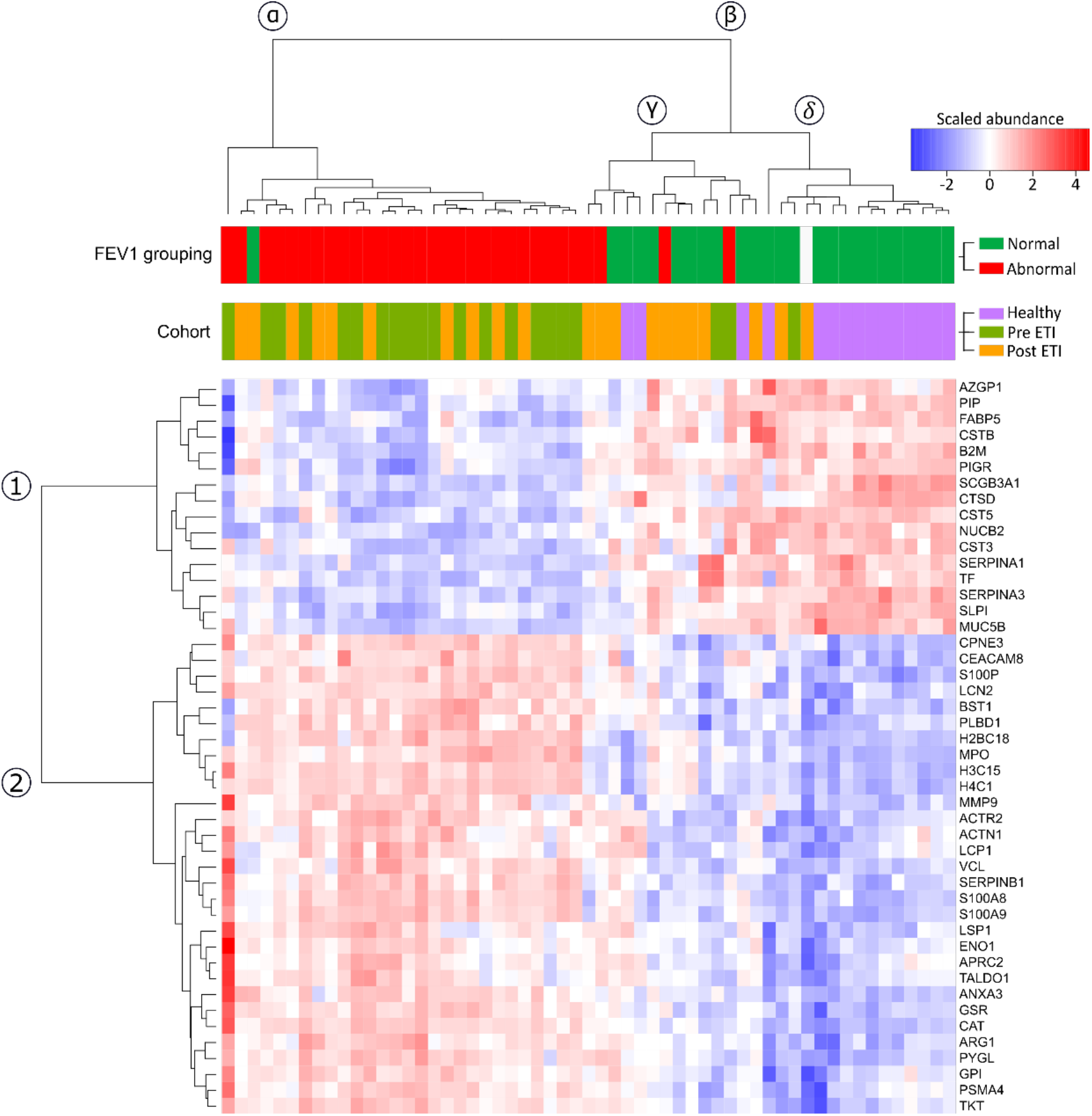
Hierarchical clustering using the proteins with the greatest discrimination between CF and healthy control samples. Healthy control, pre- and post-ETI sputum samples were clustered and displayed in a heatmap, onto which is projected the scaled abundance for the top 10% proteins that contributed to PC1 separation, clinical cohort and FEV_1_ grouping. The clusters are labelled for proteins (clades 1 and 2) and subjects (clades α, β,γ and δ. D*efinition of abbreviations:* AZGP1 = Zinc-alpha-2-glycoprotein, PIP = Prolactin-inducible protein, FABP5 = Fatty acid-binding protein 5, CSTB = Cystatin-B, B2M = Beta-2-microglobulin, PIGR = Polymeric immunoglobulin receptor, SCGB3A1 = Secretoglobin family 3A member 1, CTSD = Cathepsin D, CST5 = Cystatin-D, NUCB2 = Nucleobindin-2, CST3 = Cystatin-C, SERPINA1 = alpha-1-antiproteinase, TF = Serotransferrin, SERPINA3 = Alpha-1-antichymotrypsin, SLPI = Antileukoproteinase, MUC5B = Mucin-5B, CPNE3 = Copine-3, CEACAM8 = Carcinoembryonic antigen-related cell adhesion molecule 8, S100P = Protein S100-P, LCN2 = Neutrophil gelatinase-associated lipocalin, BST1 = ADP-ribosyl cyclase/cyclic ADP-ribose hydrolase 2, ADP-ribosyl cyclase/cyclic ADP-ribose hydrolase 2, PLBD1 = Phospholipase B-like-1, H2BC18 = Histone H2B type 2-F, MPO = Myeloperoxidase, H3C15 = Histone H3.2, H4C1 = Histone H4, MMP9 = Matrix metalloproteinase-9, ACTR2 = Actin-related protein 2, ACTN1 = Alpha-actinin-1, LCP1 = Plasyin-2, VCL = Vinculin, SERPINB1 = Leukocyte elastase inhibitor, S100A8 = Protein S100-A8, S100A9 = Protein S100-A9, LSP1 = Lymphocyte-specific protein 1, ENO1 = Alpha-enolase, ARPC2 = Actin-related protein 2/3 complex subunit 2, TALDO1 = Transaldolase, ANXA3 = Annexin-3, GSR = Glutathione reductase, CAT = Catalase, ARG1 = Arginase-1, PYGL = Glycogen phosphorylase, GPI = Glucose-6-phosphate isomerase, PSMA4 = Proteasome subunit alpha type-4 and TKT = Transketolase.

### ETI therapy is associated with reduced neutrophilic inflammation and restoration of anti-inflammatory mechanisms

Protein set enrichment analysis revealed that the proteome changes observed on PCA are largely related to neutrophil activity, in keeping with ETI reducing but not completely resolving inflammation (Figures 1e and 1f). These improvements were defined in greater detail within the hierarchical clustering (Figure 2). The proteins that contributed most to PC1 separated into two distinct groups: a pro-inflammatory set, predominantly neutrophil-derived (e.g., myeloperoxidase (MPO) and S100-A8/A9), and an anti-inflammatory set that included antiproteases predominantly derived from epithelial cells and macrophages, such as Cystatin C, Cystatin D, Alpha-1 antitrypsin, Alpha-1 antichymotrypsin and secretory leukocyte peptidase inhibitor. Those with CF and impaired lung function had a greater imbalance of pro-and ant-inflammatory proteins. This imbalance was reduced with ETI.

The protein-level data revealed 110 sputum proteins that changed (adjusted p <0.05) with ETI therapy (Figure 3a); most were increased (80/110, 72.7 %). Functional analysis confirmed that these changes were associated with reduced neutrophil activity (Figure 3b). However, we also identified changes in immunomodulatory and counter-regulatory molecular functions such as antioxidant activity, antiprotease activity and phospholipase inhibition (Figure 3c). Increased abundances were noted specifically for cystatins (cystatin B and kininogen), peroxiredoxin 5 and 6, tissue inhibitor of metalloproteinase 1 and antileukoproteinase (Supplementary table E2). Although less numerous, some neutrophil-derived proteins reduced post-ETI, including protein S100 A8/A9 (calprotectin). Figure 4 illustrates the changes seen in neutrophil mediated immunity proteins that changed the most with ETI. Post-ETI most remained different (adjusted p<0.05) from healthy controls.

**Figure 3.**
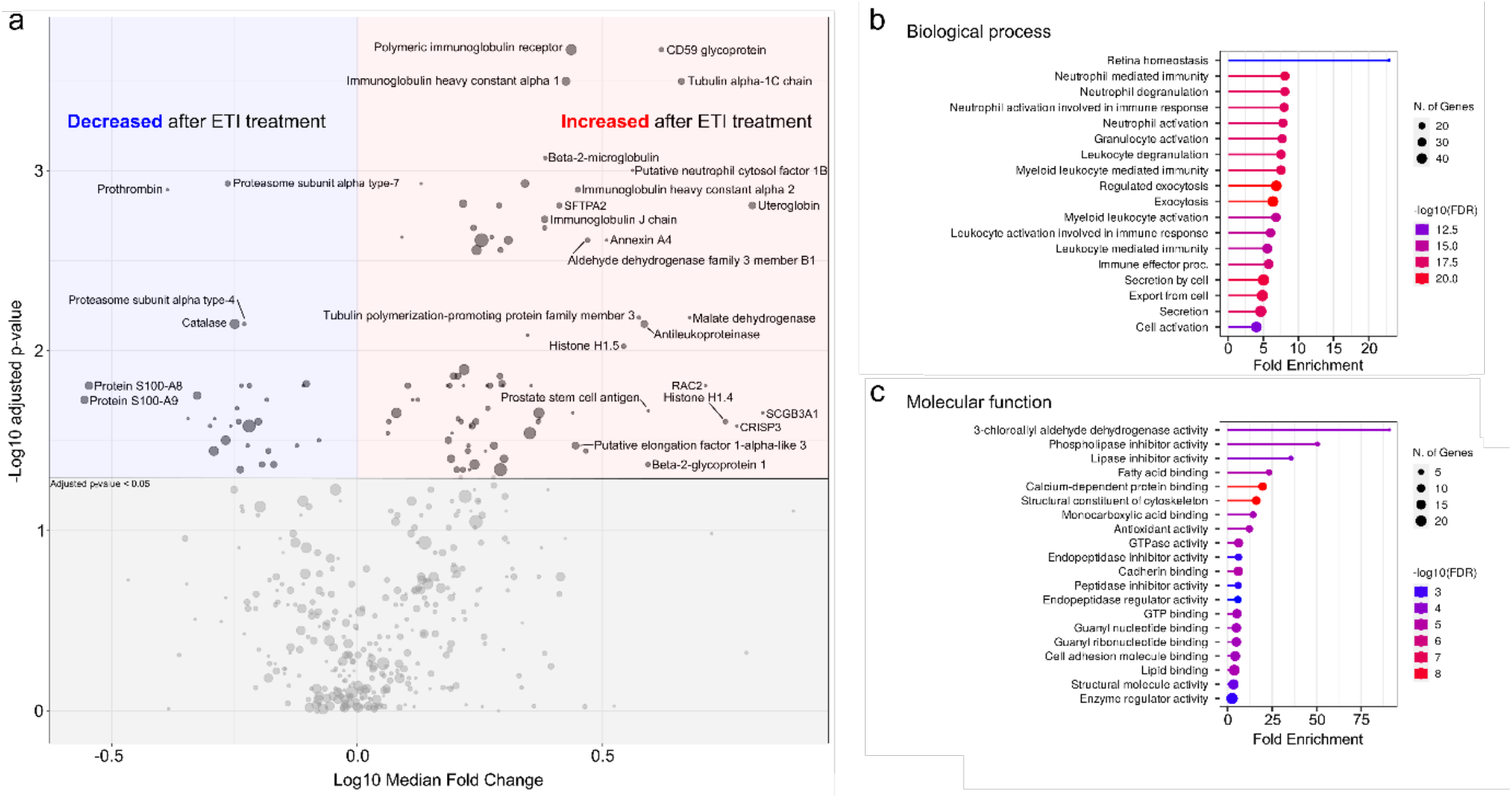
Quantitative profiling of cystic fibrosis sputum proteomes pre- and post-elexacaftor/tezacaftor/ivacaftor therapy. (a) Volcano plot displaying log10median fold change in protein abundance against adjusted p-values derived from Wilcoxon signed-rank test. Symbols corresponding to proteins with the greatest statistical confidence and highest fold change are labelled. The symbol size maps to the number of unique peptides used for quantification with the smallest being two unique peptides. The top enriched (b) biological processes and (c) molecular functions based on proteins with an adjusted p-value < 0.05.

**Figure 4.**
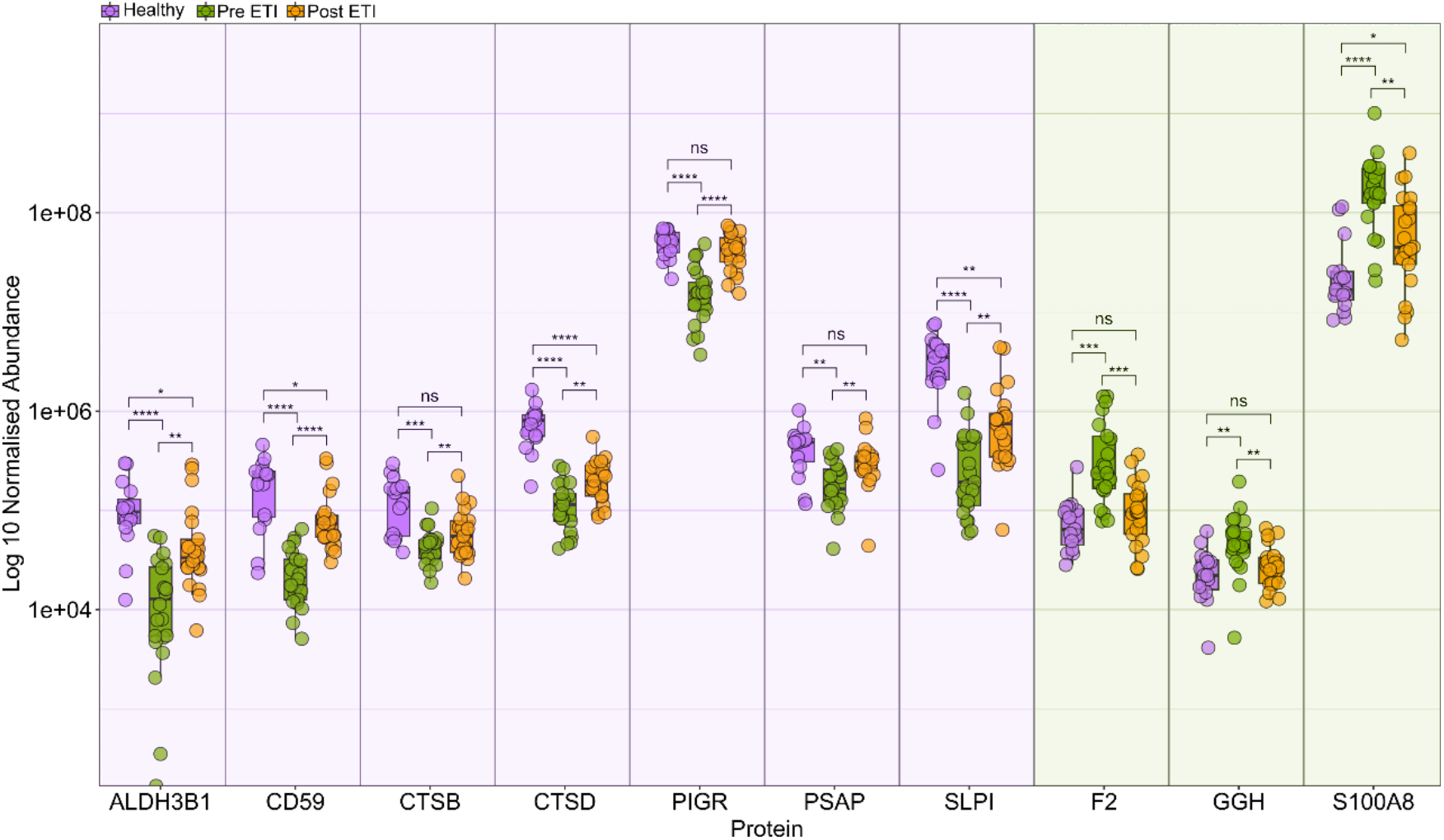
Abundance profiles for the key altered neutrophil proteins. For the ten neutrophil mediated immunity proteins with the greatest significant difference based on adjusted p-value between pre- and post-elexacaftor/ tezacaftor/ivacaftor (ETI), the normalised protein abundances are plotted, with CF subjects stratified according to whether they are pre-or post-ETI therapy. For each group, individual protein abundances, together with median and interquartile ranges are plotted. **** = p < 0.0001, *** = p < 0.001, ** = P < 0.01, * = p < 0.05, ns = not significant. *Definition of abbreviations:* ALDH3B1 = Aldehyde dehydrogenase family 3 member B1, CD59 = CD59 glycoprotein, CSTB = Cystatin-B, CTSD = Cathepsin D, PIGR = Polymeric immunoglobulin receptor, PSAP = Prosaposin, SLPI = Antileukoproteinase, F2 = Prothrombin, GGH = Gamma-glutamyl hydrolase and S100A8 = Protein S100-A8.

### Some respiratory proteases appear unchanged with CFTR modulation therapy

We performed an exploratory analysis to identify high confidence proteins which don’t appear to respond to ETI therapy. We focussed on 62 proteins with clear differential expression between pre-ETI CF and healthy controls, defined as statistically significant (adjusted p value <0.05) and at least a log2 median fold difference. We reduced this to 29 by excluding proteins whose expression changed following ETI therapy (unadjusted p value <0.05). Further refinement using the TOST procedure for equivalence testing, identified 6 proteins (cathepsin G, proteinase 3, matrix metalloproteinase 9, integrin beta-2, glycogen phosphorylase (liver form) and peptidoglycan recognition protein 1) that were unchanged by ETI therapy in CF subjects (adjusted p <0.05) (Figure 5).

**Figure 5.**
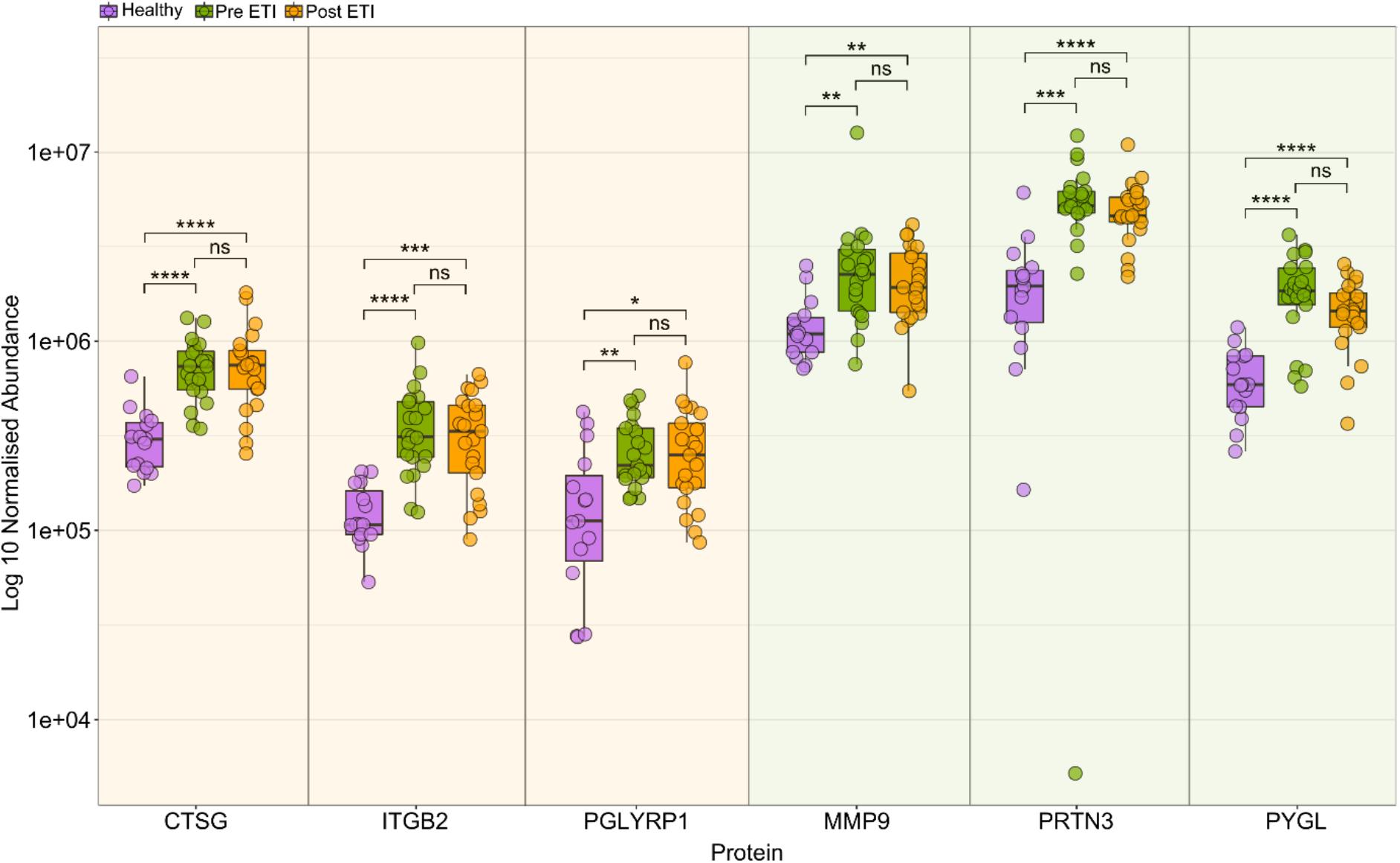
Abundance profiles for proteins minimally altered by ETI therapy. For six proteins that were minimally altered by ETI therapy based on the equivalence testing between pre- and post-elexacaftor/ tezacaftor/ivacaftor (ETI), the normalised protein abundances are plotted, with CF subjects stratified according to whether they are pre-or post-therapy. For each group, individual protein abundances, together with median and interquartile ranges are plotted. **** = p < 0.0001, *** = p < 0.001, ** = P < 0.01, * = p < 0.05, ns = not significant. *Definition of abbreviations:* CTSG = Cathepsin G, PRTN3 = Myeloblastin, PGLYRP1 = Peptidoglycan recognition protein 1, ITGB2 = Integrin beta-2, PYGL = Glycogen phosphorylase (liver) and MMP9 = Matrix metalloproteinase-9.

## DISCUSSION

In this study, we describe for the first time the effect of ETI on CF airway pathology using sputum proteomics. Compared with healthy controls the proteome pre-ETI was characterised by excessive neutrophil-derived proteins and a reduction in proteins capable of mitigating the toxic effects of neutrophils on the airway. With ETI therapy this imbalance improved but for most CF subjects remained distinct from healthy controls, even when normal lung function was achieved. Notably, a far greater number of proteins increased than decreased with therapy, many of these possessing anti-inflammatory properties. Although some neutrophil-derived proteins reduced with ETI, several important respiratory proteases showed little difference.

Currently CF lung disease is monitored clinically by tracking FEV_1_%. As previously shown, the proteome differences between CF and healthy controls in this study become more pronounced with progressive lung function impairment [10]. However, despite significant improvements in both lung function and sputum proteome these appear largely independent, suggesting we are measuring separate elements of CF lung pathophysiology. This was not unanticipated as spirometric measures are insensitive for early lung disease in comparison to radiology and lung clearance index [23]. Importantly, the post-ETI proteome differed from both pre-ETI and healthy controls, appearing to adopt an intermediate state consistent with incomplete resolution of the dysregulated neutrophilic inflammation central to CF lung disease. This intermediate state was observed even in subjects achieving normal lung function with ETI therapy confirming that spirometry may be insensitive for capturing residual disease activity and complementary biomarkers could be beneficial.

Our data suggest that CFTR modulation creates a less immunogenic airway environment, however the exact mechanisms behind this needs to be further elucidated. It is likely there is a direct immunological effect by influencing the behaviour of immune cells carrying CFTR protein, but also there is also increasing evidence of a reduction in microbial burden which may influence the inflammatory mileu [24-26]. Within this intermediate state there was partial recovery of a range of regulatory proteins that mitigate the toxic effects of dysregulated neutrophil responses. These included proteins controlling the oxidative stress and proteolysis caused by the release of NETs and various secretory proteins. Many more proteins increased than decreased with therapy (80 vs 30) highlighting the significance of the anti-inflammatory as well as pro-inflammatory processes and how their balance determines homeostasis. There were a range of different antiproteases, antioxidants and immunomodulatory proteins. One prominent novel immunomodulatory example was CD59, a complement regulatory protein that inhibits membrane attack complex formation [27]. The Pre-ETI levels of CD59 were significantly lower than in healthy controls and then increased with therapy. In contrast, higher levels of CD59 are associated with myocardial infarction, pancreatitis, and sepsis, and appear to predict subsequent risk of allograft dysfunction in lung transplant recipients [28-31]. As such its role in the CF airway is unclear at this point. Uteroglobin is another immunomodulatory protein with a range of effects on innate and adaptive responses [32]. Deficits have been described in CF, alongside negative correlations with airway inflammation and disease severity [33-35]. We identified reduced levels of uteroglobin which improved with therapy.

Despite some neutrophil proteins being reduced with ETI, such as S100A8/A9 (calprotectin), we observed little alteration in important respiratory proteases; proteinase 3, cathepsin G and matrix metalloproteinase 9. Proteinase 3 and cathepsin G, along with neutrophil elastase, are serine proteases released by neutrophils. These are in a physiological balance with antiproteases, with members of the SERPIN family being particularly important including alpha-1 antitrypsin. An imbalance in favour of these proteases results in breakdown of the lung matrix and is thought to result in the progressive destructive bronchiectasis observed in CF [36]. Its continued presence highlights the potential for progressive lung disease in those on ETI therapy. Matrix metalloproteinase 9 is secreted by neutrophils as well as epithelial cells. It appears to indirectly cause lung destruction by several methods, including inactivating alpha-1 antitrypsin [37]. These findings likely reveal that CFTR modulation therapy alters a complex inflammatory environment in the CF lung which is important to understand when we evaluate long-term outcomes with these therapies. However, as structural lung damage and bronchiectasis become established these changes contribute to disease mechanisms potentially independent of CFTR activity, such as through airway infection as seen in other causes of bronchiectasis. In addition, disease activity appears characterised by imbalances we observed in protein activity originating from both immune and epithelial cells, and these may not respond uniformly to ETI. As such, it may be difficult to predict the effect of interventions on this complex network of disease mechanisms thus justifying the use of untargeted approaches, such as sputum proteomics, to evaluate these responses without assumptions.

Reduced levels of airway neutrophils and inflammatory cytokines have been seen previously with CFTR modulation, but not consistently in all studies [18, 19, 38]. As CF is a multisystem disease inflammatory changes are not necessarily specific to the lung. Our results are also in accordance with studies showing that CFTR modulation reduces systemic inflammation with lower levels of circulating blood neutrophils and pro-inflammatory cytokines, such as IL-1β, IL6, IL8, IL-17A, IL18 and TNFα [14-17]. It should be noted that studies examining systemic and local cytokine levels mainly reflect the signals responsible for neutrophil recruitment and activation within the airway. In contrast, our proteome data provide details of the airway environment that is created by excessive airway neutrophils, the subsequent predominantly epithelial responses, and the influence of CFTR modulation on both.

There are several limitations with this study. This is a single-centre study and therefore our findings need replicating in a broader population to confirm their generalisability. It was not feasible to study those with severe lung disease as they were commenced on ETI therapy immediately prior to licensing in the UK on a named patient basis which predated the commencement of this study. Otherwise, our patients are broadly typical of adult CF patients. It is notable that we were unable to obtain samples on a number of patients post therapy due to reduced sputum loads so it is unclear whether these patients may have differing proteome responses. We acknowledge the potential impact of subjects where the pre-ETI samples were taken whilst taking dual CFTR modulators and variations in the time a subject received ETI before repeat sampling, but these potential confounding effects were examined, and we found no obvious effect on the sputum proteome.

In conclusion, our analyses support the concept that sputum proteomics can provide insights into to the processes that directly result in CF lung destruction and how they are modified by therapeutic intervention, in this case using ETI. This study identified imbalances in pro- and anti-inflammatory proteins that shift with therapy toward an intermediate state that is not fully represented by lung function status. It is a possible that false reassurance is provided by achieving dramatic improvements and even normalisation of lung function post-ETI. Our data support a post-ETI intermediate state that has the potential for ongoing airway damage and therefore its relevance to clinical outcomes needs to be established.

## Data Availability

All data produced in the present study are available upon reasonable request to the authors

## ACKNOWLEDGEMENTS

This report is independent research supported by the North West Lung Centre Charity at Manchester University NHS Foundation Trust. The views expressed in this publication are those of the author(s) and not necessarily those of the NHS, the North West Lung Centre Charity or the Department of Health. The authors would like to acknowledge the Manchester Allergy, Respiratory and Thoracic Surgery Biobank, the North West Lung Centre Charity and the National Institute for Health Research (NIHR) Manchester Clinical Research Facility for supporting this project. In addition, we would to thank the study participants for their contribution, as well as the medical and nursing staff especially Jo Hyde, Cassandra McNaughton and Daniel Tewkesbury at the Manchester Adult CF Centre for their invaluable assistance.

