## Supplementary material for "The influence of highly effective modulator therapies on the sputum proteome in cystic fibrosis"

**Supplementary methods**

***Sputum processing***

Sputum samples were thawed and weighed after which four times the weight of 0.1% (w/v) (6.5mM) dithiothreitol (DTT), dissolved in 0.01 M phosphate-buffered saline, was added. The mixture was vortexed vigorously for 30 s before incubating for 30 minutes on a roller mixer at room temperature. The samples were vortexed for 30 s every 10 minutes during this time. Samples were centrifuged at 450 g at 4 °C for 10 minutes and the supernatant fraction was transferred to low protein binding microfuge tubes and re-centrifuged at 16,000 g for 30 minutes at 4 °C. The liquid fractions were removed and combined. Processed sputum samples were aliquoted and stored at -80 °C.

***Proteomics***

Sputum samples were diluted in 25 mM ammonium bicarbonate (AmBic) to a final volume of 160 µL where samples had a total protein concentration of 0.16 µg/uL. Samples were treated with a 1% (w/v) solution of RapiGestTM SF Surfacant (Waters) to a final concentration of 0.05% (w/v) before incubation at 80 °C for 10 minutes. Disulfide bonds were reduced by the addition of DTT during sputum processing, so an additional 10 µL of 25mM AmBic was added. Iodoacetamide (IA) was added to a final concentration of 9 mM to alkylate the free sulfhydryl groups on the cysteine residues. After incubation for 30 minutes at room temperature in the dark trypsin was added to a 50:1 protein: protease ratio and incubated overnight at 37 °C. Digestion was stopped by the addition of trifluoroacetic acid (TFA) to a final concentration of 0.5% (v/v) and incubated for 45 minutes at 37 °C. The samples were centrifuged to remove particulates for 15 minutes up to 13,000 g. Off-line desalting was performed using C18 trap columns, (Evotips, Evosep) as per the manufactures protocol. Peptides were eluted using acetonitrile 60% (v/v) with 0.1% (v/v) formic acid (FA), dried down using a SpeedVac and suspended in 200 µL 973. The samples were then subject to LC-MS/MS analysis. Peptides were analysed by LC-MS/MS using an Ultimate 3000 nano system (Dionex/Thermo Fisher Scientific, Hemel Hempstead, UK) coupled to QExactive-HF mass spectrometer (Thermo Fisher Scientific, Hemel Hempstead, UK) to aquire the masses of peptides derived from proteins extracted from the sputum samples. Peptides were loaded onto a trap column (50cm) (Acclaim PepMap 100) at 12 µL/min over 7 min with an aqueous solution containing 0.1% (v/v) TFA and 2% (v/v) MeCN. The trap column was set in-line with an analytical column (Easy-Spray PepMap® RSLC) (Dionex). Peptides were eluted by applying a linear gradient of 3.8% to 50% solvent B (acetonitrile 80% (v/v) with 0.1% (v/v) FA) over 35 min at 300 nL/min. Solvent A was HPLC grade water with 0.1% (v/v) formic acid. The mass spectrometer was operated in data dependent positive (ESI+) mode and full scan MS spectra (300-2000 m/z) were acquired in the Orbitrap. The 16 most intense multiply charged ions ($z \geq 2$) were sequentially isolated and fragmented in the octopole collision cell by high energy collisional dissociation (HCD) and detected in the Orbitrap.

***Protein analysis***

The total protein concentration of sputum varied considerably, samples were therefore normalised to a fixed total protein concentration prior to alkylation and tryptic digestion. Data analysis including run alignment and peak picking was carried out in Progenesis QI for Proteomics v4 (Nonlinear Dynamics), while database searching was carried out by MASCOT search engine (Matrixscience version 2.6). Samples were searched against a database comprised of all reviewed human sequences in the UniProt Human database (20,356 sequences; updated 12/08/2020) using trypsin as the specified enzyme, carbamidomethylation of cysteine as fixed modification, methionine oxidation as variable modification and one trypsin missed cleavage, precursor and fragment ion error tolerances were set to 10 ppm and 0.01 Da, respectively. The false discovery rate (FDR) was calculated using the decoy database tool in MASCOT. Only those proteins identified with an FDR <1% were accepted.

**Table E1 Individual clinical and demographic characteristics of CF participants**

| ID | Stratum | Time on therapy at repeat sampling | Baseline modulator | Gender | Gene1 | Gene2 | Pancreatic status | Ps | BCC | Baseline FEV1 | Baseline FEV1% | V2 FEV 1 | V2 FEV1 % |
|---|---|---|---|---|---|---|---|---|---|---|---|---|---|
| 2227 | ETI | 56 | Tez/Iva | F | F508del | F508del | Insufficient | Yes | Yes | 2.34 | 64 | 2.74 | 75 |
| 2420 | ETI | 130 | None | F | F508del | R792X | Insufficient | Yes | No | 1.7 | 61 | 2.05 | 74 |
| 2511 | ETI | 39 | Lum/Iva | M | F508del | F508del | Insufficient | Yes | No | 2.01 | 52 | 2.5 | 64 |
| 2833 | ETI | 220 | Tez/Iva | F | F508del | F508del | Insufficient | Yes | Yes | 1.26 | 46 | 1.28 | 47 |
| 3125 | ETI | 235 | Tez/Iva | M | F508del | F508del | Insufficient | Yes | No | 2.77 | 67 | 2.92 | 70 |
| 3452 | ETI | 254 | None | M | F508del | 1154insTC | Insufficient | Yes | No | 1.9 | 44 | 2.51 | 59 |
| 4763 | ETI | 93 | None | M | F508del | 2711delT | Insufficient | No | Yes | 2.35 | 58 | 3.08 | 76 |
| 5076 | ETI | 189 | None | F | F508del | 17171G>A | Insufficient | Yes | No | 3.09 | 89 | 3.68 | 106 |
| 5117 | ETI | 246 | Tez/Iva | M | F508del | F508del | Insufficient | Yes | Yes | 2.77 | 68 | 3.61 | 90 |
| 5268 | ETI | 252 | Tez/Iva | M | F508del | F508del | Insufficient | Yes | No | 2.19 | 58 | 2.81 | 75 |
| 5270 | ETI | 161 | Tez/Iva | F | F508del | F508del | Insufficient | Yes | No | 1.79 | 59 | 1.67 | 55 |
| 5369 | ETI | 103 | Tez/Iva | F | F508del | F508del | Insufficient | Yes | No | 3.59 | 102 | 3.93 | 112 |
| 6013 | ETI | 189 | Lum/Iva | F | F508del | F508del | Insufficient | Yes | No | 2.62 | 77 | 3.06 | 91 |
| 6018 | ETI | 49 | None | F | F508del | R553X | Insufficient | Yes | No | 1.84 | 65 | 2.49 | 88 |
| 6021 | ETI | 194 | None | F | F508del | 3659delC | Insufficient | Yes | No | 1.6 | 54 | 1.8 | 61 |
| 6054 | ETI | 139 | None | M | F508del | 2184delA | Insufficient | Yes | No | 1.35 | 33 | 1.9 | 47 |
| 6063 | ETI | 259 | Tez/Iva | M | F508del | F508del | Insufficient | Yes | No | 2.46 | 63 | 3.27 | 85 |
| 6067 | ETI | 310 | Tez/Iva | M | F508del | F508del | Insufficient | Yes | No | 2.52 | 54 | 2.94 | 63 |
| 6092 | ETI | 92 | Tez/Iva | F | F508del | F508del | Insufficient | Yes | No | 2.64 | 77 | 3.44 | 100 |
| 6127 | ETI | 128 | None | M | F508del | W1098R | Insufficient | Yes | No | 3.25 | 74 | 4.39 | 100 |
| 6131 | ETI | 79 | None | M | F508del | F508del | Insufficient | Yes | No | 3.19 | 76 | No data | No data |
| 2541 | Control | N/A | None | M | 1154ins TC | 2118delAA CT | Insufficient | Yes | No | 2.56 | 74 | 2.59 | 75 |
| 2587 | Control | N/A | None | M | I507del | 2711delT | Insufficient | Yes | No | 2.37 | 66 | 2.5 | 69 |

| ID | Stratum | Time on therapy at repeat sampling | Baseline modulator | Gender | Gene1 | Gene2 | Pancreatic status | Ps | BCC | Baseline FEV1 | Baseline FEV1% | V2 FEV 1 | V2 FEV1 % |
|---|---|---|---|---|---|---|---|---|---|---|---|---|---|
| 2896 | Control | N/A | None | M | R334W | R75X | Sufficient | No | No | 3.74 | 72 | 3.5 | 68 |
| 2937 | Control | N/A | None | M | L218X | L218X | Insufficient | Yes | No | 2.75 | 71 | 2.86 | 75 |
| 3257 | Control | N/A | None | F | R553X | Q493X | Insufficient | Yes | No | 1.48 | 50 | 1.63 | 56 |
| 6303 | Control | N/A | None | M | R553X | R1162X | Insufficient | No | No | 1.75 | 40 | 1.54 | 35 |

Abbreviations: Percent predicted FEV1 (FEV1%), M (male), F(female), ivacaftor (iva), lumacaftor (lum), tezacaftor (tez)

Fully anonymised Study IDs were provided that cannot be identified outside the research group.

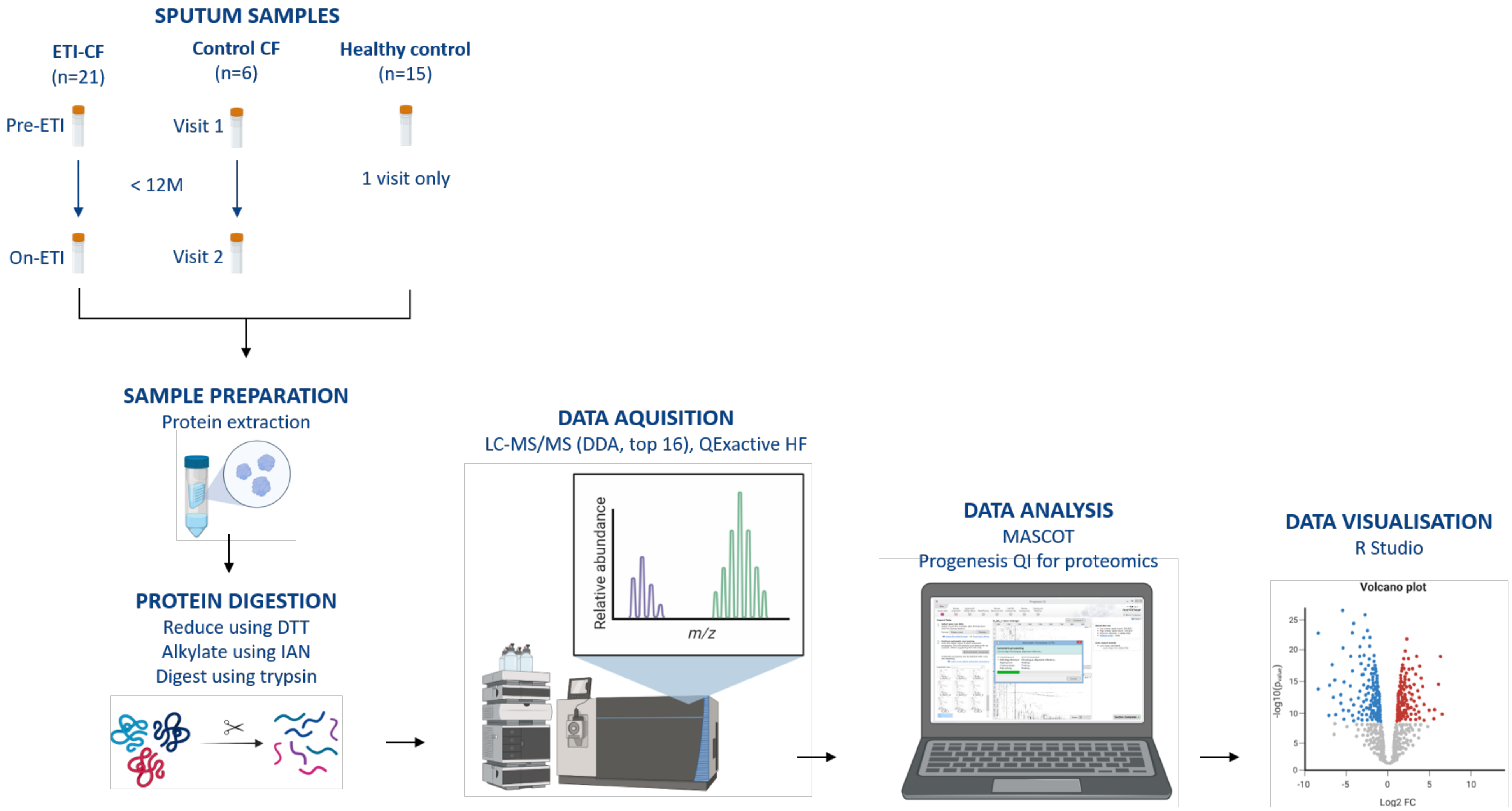


**Supplementary Figure 1. Workflow diagram.**

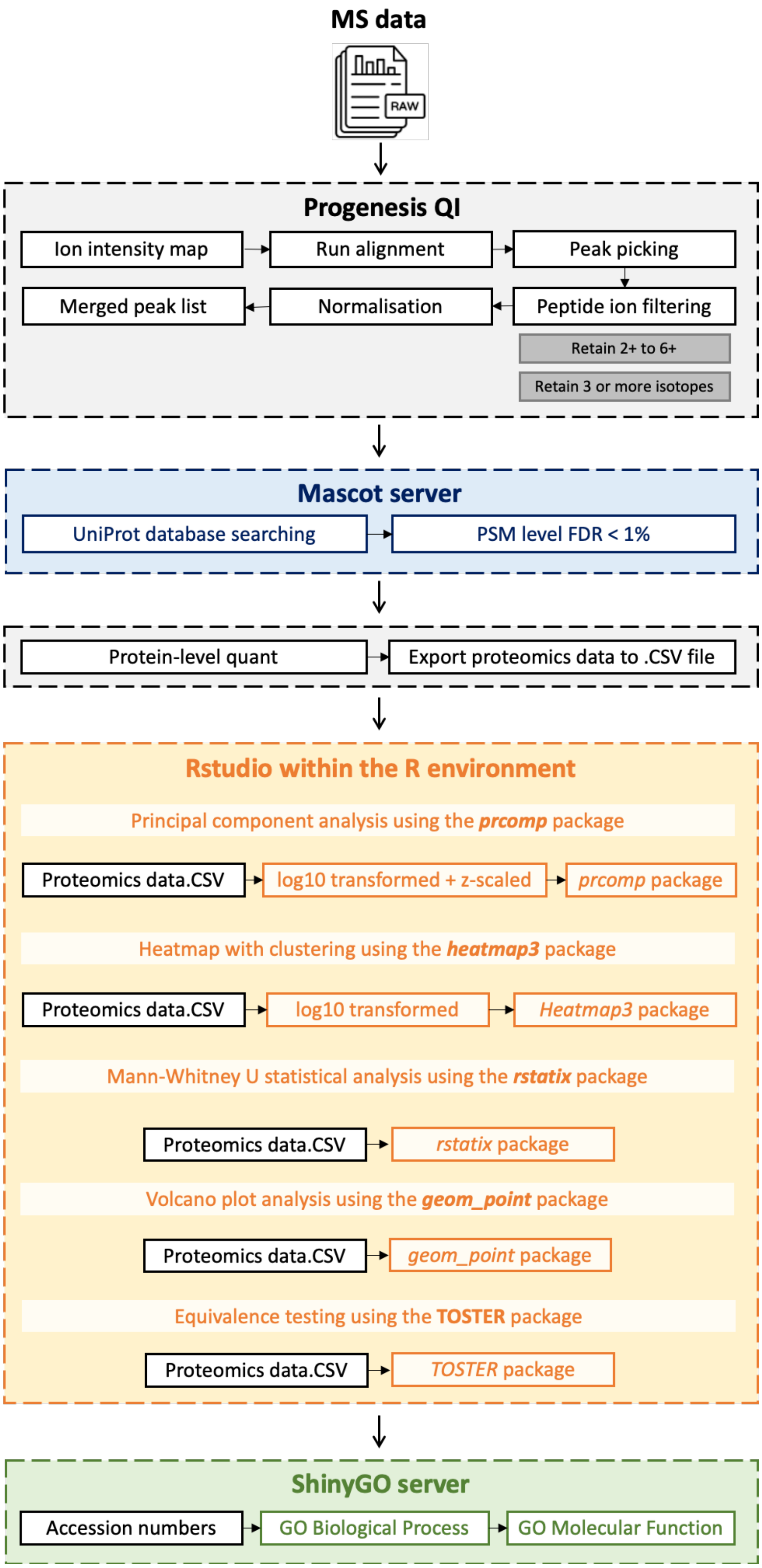


**Supplementary Figure 2. Bioinformatics workflow.**

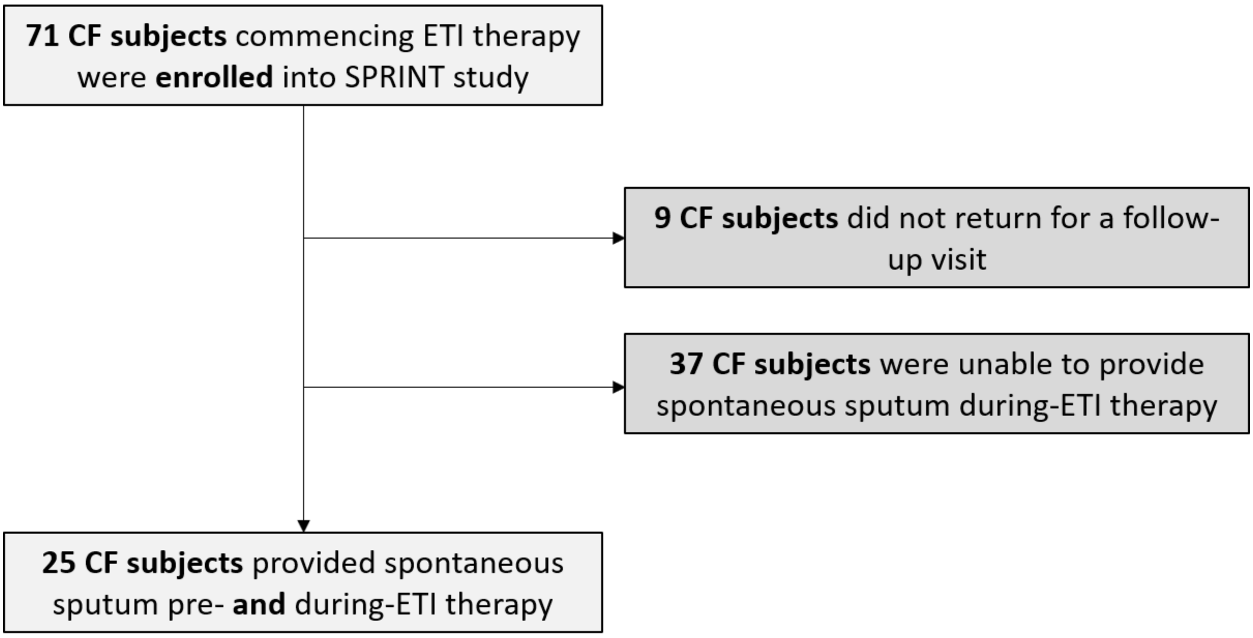


**Supplementary Figure 3, Consort diagram for recruited patients**

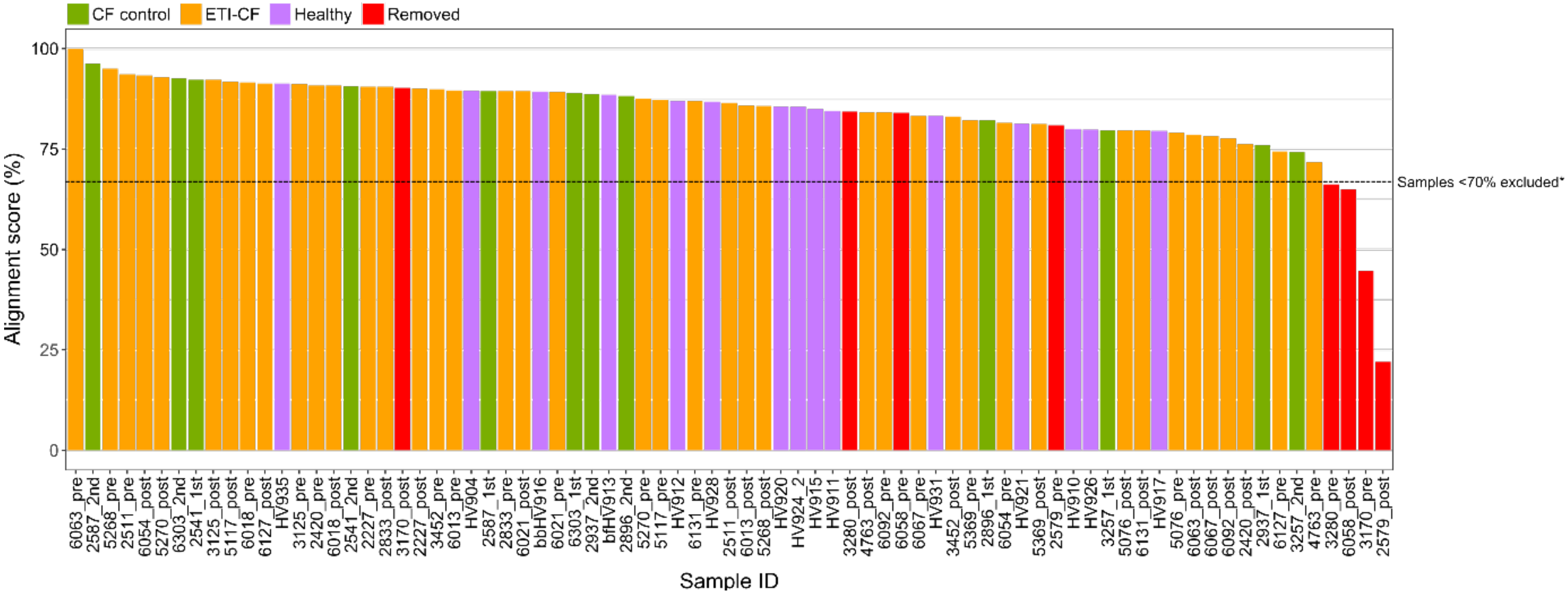


**Supplementary Figure 4. Sample Alignment.**

For each sample Progenesis QI performed automatic sample alignment, providing an alignment score presented as a percentage of matchable peaks that are matched to the reference run (6063_pre). The bar chart above presents the alignment score for each sample in the analysis, colour coded based on cohort; CF control (green), ETI-CF (orange) and healthy (purple). Samples with an alignment score of < 70%, *along with their paired sample (i.e. both pre and post ETI) were removed from the analysis and are highlighted in red.

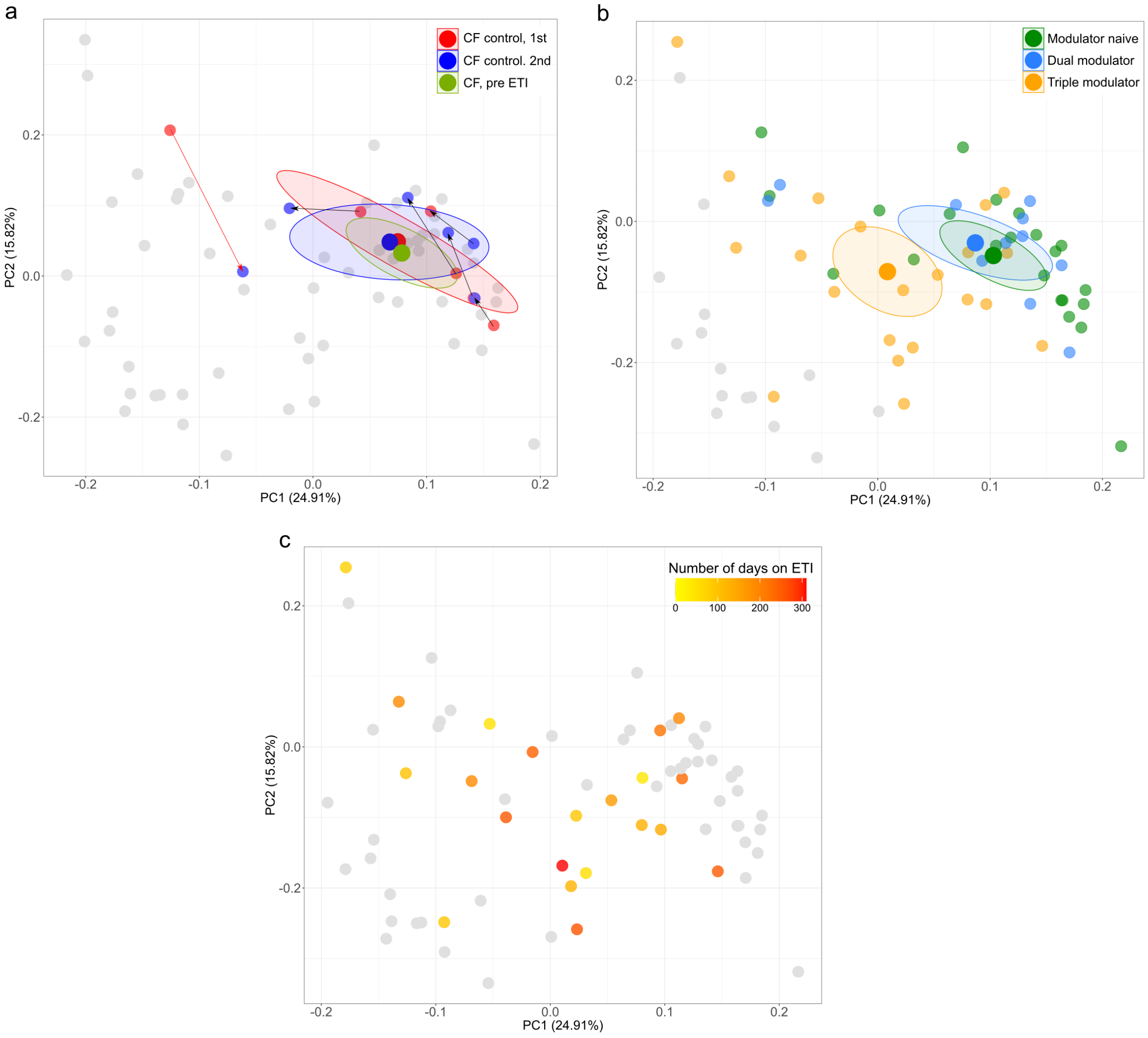


**Supplementary Figure 5. Principal component analysis of sputum protein profiles.** Principal component analysis of 446 core sputum proteins. A centroid marker (large dot) with 95 % confidence ellipse was calculated where appropriate. (A) PCA of longitudinal CF controls not eligible for ETI. Lines indicate movement of paired samples. (B) Plot of baseline modulator status. (C) PCA colour coded by the number of days on ETI.

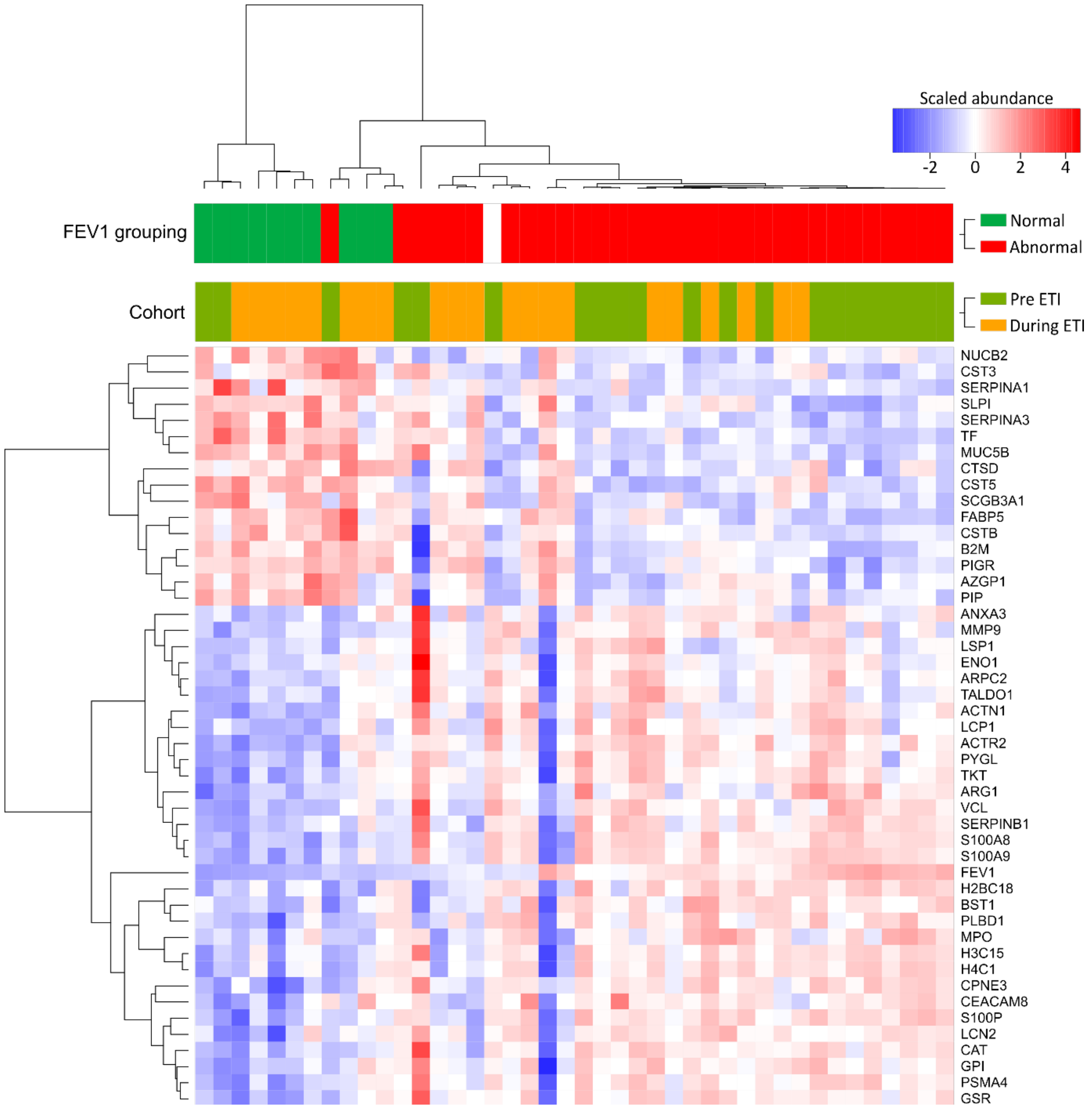


**Supplementary Figure 6. Hierarchical clustering using the proteins with the greatest discrimination between pre and post ETI (PC1).** Pre- and post- ETI sputum samples were clustered and displayed in a heatmap, onto which is projected the scaled abundance for the top 10% proteins that contributed to PC1 separation, clinical cohort and FEV1 grouping. D*efinition of abbreviations:* AZGP1 = Zinc-alpha-2-glycoprotein, PIP = Prolactin-inducible protein, FABP5 = Fatty acid-binding protein 5, CSTB = Cystatin-B, B2M = Beta-2-microglobulin, PIGR = Polymeric immunoglobulin receptor, SCGB3A1 = Secretoglobin family 3A member 1, CTSD = Cathepsin D, CST5 =  Cystatin-D ,NUCB2 = Nucleobindin-2, CST3 = Cystatin-C, SERPINA1 = alpha-1-antiproteinase, TF = Serotransferrin, SERPINA3 = Alpha-1-antichymotrypsin, SLPI = Antileukoproteinase, MUC5B = Mucin-5B, CPNE3 = Copine-3, CEACAM8 = Carcinoembryonic antigen-related cell adhesion molecule 8, S100P = Protein S100-P, LCN2 = Neutrophil gelatinase-associated lipocalin, BST1 = ADP-ribosyl cyclase/cyclic ADP-ribose hydrolase 2, ADP-ribosyl cyclase/cyclic ADP-ribose hydrolase 2, PLBD1 = Phospholipase B-like-1, H2BC18 = Histone H2B type 2-F, MPO = Myeloperoxidase, H3C15 = Histone H3.2, H4C1 = Histone H4, MMP9 =

Matrix metalloproteinase-9, ACTR2 = Actin-related protein 2, ACTN1 = Alpha-actinin-1, LCP1 = Plasyin-2, VCL = Vinculin, SERPINB1 = Leukocyte elastase inhibitor, S100A8 = Protein S100-A8 , S100A9 = Protein S100-A9, LSP1 = Lymphocyte-specific protein 1, ENO1 = Alpha-enolase, ARPC2 = Actin-related protein 2/3 complex subunit 2, TALDO1 = Transaldolase, ANXA3 = Annexin-3, GSR = Glutathione reductase, CAT = Catalase, ARG1 = Arginase-1, PYGL = Glycogen phosphorylase, GPI = Glucose-6-phosphate isomerase, PSMA4 = Proteasome subunit alpha type-4 and TKT = Transketolase.

**Table E2 Proteins showing a significant correlation (unadjusted p value <0.05) using Mann Whitney**

| Protein | Post | Pre | fold change | p | p.adj |
|---|---|---|---|---|---|
| Polymeric immunoglobulin receptor | 42714431.1 | 15641786.9 | 2.73 | 9.54 E-07 | 2.13 E-04 |
| CD59 glycoprotein | 73405.7267 | 17603.8058 | 4.17 | 9.54 E-07 | 2.13 E-04 |
| Immunoglobulin heavy constant alpha 1 | 117656452 | 44164866.7 | 2.66 | 2.86 E-06 | 3.19 E-04 |
| Tubulin alpha-1C chain | 106334.742 | 23208.106 | 4.58 | 2.86 E-06 | 3.19 E-04 |
| Beta-2-microglobulin | 599853.014 | 248148.183 | 2.42 | 9.54 E-06 | 8.51 E-04 |
| Putative neutrophil cytosol factor 1B | 19615.6933 | 5384.65753 | 3.64 | 1.34 E-05 | 9.96 E-04 |
| Proteasome subunit alpha type-7 | 229256.079 | 421184.369 | 0.54 | 2.38 E-05 | 1.18 E-03 |
| Immunoglobulin lambda-like polypeptide 5 | 7656088.63 | 5675504.27 | 1.35 | 2.38 E-05 | 1.18 E-03 |
| Complement factor B | 633645.603 | 288331.268 | 2.20 | 2.38 E-05 | 1.18 E-03 |
| Prothrombin | 97008.7514 | 236422.96 | 0.41 | 3.15 E-05 | 1.28 E-03 |
| Immunoglobulin heavy constant alpha 2 | 16395006.5 | 5819626.74 | 2.82 | 3.15 E-05 | 1.28 E-03 |
| Galectin-3-binding protein | 771924.142 | 469585.214 | 1.64 | 4.10 E-05 | 1.52 E-03 |
| Heterogeneous nuclear ribonucleoproteins A2/B1 | 89141.9881 | 45792.7761 | 1.95 | 5.25 E-05 | 1.56 E-03 |
| Pulmonary surfactant-associated protein A2 | 46756.6686 | 18111.8572 | 2.58 | 5.25 E-05 | 1.56 E-03 |

| | | | | | |
|---|---|---|---|---|---|
| **Uteroglobin** | 2310884.16 | 361589.293 | 6.39 | 5.25E-05 | 1.56E-03 |
| **Immunoglobulin J chain** | 11858690.7 | 4919357.1 | 2.41 | 6.68E-05 | 1.86E-03 |
| **Immunoglobulin kappa constant** | 100883708 | 58418178.3 | 1.73 | 8.39E-05 | 2.08E-03 |
| **Fatty acid-binding protein 5** | 112991.125 | 46880.7941 | 2.41 | 8.39E-05 | 2.08E-03 |
| **Immunoglobulin kappa variable 3-20** | 355917.061 | 288344 | 1.23 | 1.05E-04 | 2.34E-03 |
| **Cystatin-B** | 794464.405 | 422443.705 | 1.88 | 1.05E-04 | 2.34E-03 |
| **Mucin-5AC** | 12687354.6 | 7077308.19 | 1.79 | 1.31E-04 | 2.43E-03 |
| **Prominin-1** | 249063.27 | 122529.635 | 2.03 | 1.31E-04 | 2.43E-03 |
| **Aldehyde dehydrogenase family 3 member B1** | 33254.2585 | 11273.979 | 2.95 | 1.31E-04 | 2.43E-03 |
| **Annexin A4** | 23629.0844 | 7327.63978 | 3.22 | 1.31E-04 | 2.43E-03 |
| **Vimentin** | 1217283.77 | 694977.583 | 1.75 | 1.61E-04 | 2.76E-03 |
| **14-3-3 protein beta/alpha** | 278799.587 | 142268.486 | 1.96 | 1.61E-04 | 2.76E-03 |
| **Cytosolic non-specific dipeptidase** | 6610.98973 | 3539.22753 | 1.87 | 4.26E-04 | 6.55E-03 |
| **Tubulin polymerization-promoting protein family member 3** | 21298.0097 | 5674.62939 | 3.75 | 4.26E-04 | 6.55E-03 |
| **Malate dehydrogenase, mitochondrial** | 11617.848 | 2439.78866 | 4.76 | 4.26E-04 | 6.55E-03 |
| **Catalase** | 2349226.96 | 4176357.33 | 0.56 | 5.10E-04 | 7.11E-03 |

| | | | | | |
|---|---|---|---|---|---|
| **Proteasome subunit alpha type-4** | 59760.6024 | 101586.909 | 0.59 | 5.10E-04 | 7.11E-03 |
| **Antileukoproteinase** | 739622.38 | 192212.815 | 3.85 | 5.10E-04 | 7.11E-03 |
| **Mucin-1** | 41809.3941 | 18792.8663 | 2.22 | 6.07E-04 | 8.20E-03 |
| **Histone H1.5** | 220514.434 | 63120.4407 | 3.49 | 7.21E-04 | 9.46E-03 |
| **BPI fold-containing family B member 1** | 20431179.6 | 12370305.8 | 1.65 | 1.00E-03 | 1.27E-02 |
| **Chloride intracellular channel protein 1** | 282768.028 | 179994.505 | 1.57 | 1.18E-03 | 1.38E-02 |
| **Ezrin** | 88782.2678 | 55372.6389 | 1.60 | 1.18E-03 | 1.38E-02 |
| **WAP four-disulfide core domain protein 2** | 2632026.69 | 1347007.52 | 1.95 | 1.18E-03 | 1.38E-02 |
| **Olfactomedin-4** | 225513.704 | 286386.038 | 0.79 | 1.37E-03 | 1.53E-02 |
| **Actin, cytoplasmic 1** | 2309445 | 1170513.34 | 1.97 | 1.37E-03 | 1.53E-02 |
| **Protein S100-A8** | 45041466 | 158688382 | 0.28 | 1.86E-03 | 1.57E-02 |
| **Protein S100-P** | 787167.793 | 1355733.44 | 0.58 | 1.60E-03 | 1.57E-02 |
| **Gamma-glutamyl hydrolase** | 26961.5437 | 44739.7131 | 0.60 | 1.60E-03 | 1.57E-02 |
| **Chitotriosidase-1** | 64800.6317 | 83470.3517 | 0.78 | 1.86E-03 | 1.57E-02 |
| **Metalloproteinase inhibitor 1** | 325896.164 | 256950.991 | 1.27 | 1.86E-03 | 1.57E-02 |
| **Apolipoprotein C-I** | 57808.4467 | 37879.7545 | 1.53 | 1.60E-03 | 1.57E-02 |

| | | | | | |
|---|---|---|---|---|---|
| **Aldo-keto reductase family 1 member A1** | 14116.2397 | 9144.147 | 1.54 | 1.86E-03 | 1.57E-02 |
| **Ras-related protein Rab-1A** | 11005.5348 | 6682.64492 | 1.65 | 1.60E-03 | 1.57E-02 |
| **Programmed cell death 6-interacting protein** | 8136.51143 | 4424.67736 | 1.84 | 1.86E-03 | 1.57E-02 |
| **Nucleobindin-2** | 78248.8278 | 41978.6819 | 1.86 | 1.60E-03 | 1.57E-02 |
| **Transgelin-2** | 97107.5898 | 49034.2365 | 1.98 | 1.86E-03 | 1.57E-02 |
| **Leucine-rich repeat flightless-interacting protein 1** | 6016.71528 | 2659.65283 | 2.26 | 1.86E-03 | 1.57E-02 |
| **Ras-related C3 botulinum toxin substrate 2** | 42564.2197 | 8293.32199 | 5.13 | 1.86E-03 | 1.57E-02 |
| **Vinculin** | 137259.205 | 291038.831 | 0.47 | 2.15E-03 | 1.78E-02 |
| **Protein S100-A9** | 72489053.1 | 260907501 | 0.28 | 2.48E-03 | 1.87E-02 |
| **Proteasome subunit alpha type-3** | 56704.1349 | 86668.7617 | 0.65 | 2.48E-03 | 1.87E-02 |
| **Immunoglobulin heavy variable 6-1** | 142403.153 | 109793.91 | 1.30 | 2.48E-03 | 1.87E-02 |
| **Immunoglobulin lambda variable 7-46** | 84691.84 | 63712.5231 | 1.33 | 2.48E-03 | 1.87E-02 |
| **C-X-C motif chemokine 17** | 1561.23042 | 0 | NA | 2.35E-03 | 1.87E-02 |
| **Copine-3** | 31401.7741 | 55215.0549 | 0.57 | 2.86E-03 | 2.09E-02 |
| **Apolipoprotein E** | 7147.1404 | 3871.40779 | 1.85 | 2.86E-03 | 2.09E-02 |
| **Prostate stem cell antigen** | 28353.1456 | 7230.41555 | 3.92 | 3.00E-03 | 2.16E-02 |

| | | | | | |
|---|---|---|---|---|---|
| **Ceruloplasmin** | 1767046.39 | 1469912.06 | 1.20 | 3.28E-03 | 2.22E-02 |
| **Alpha-amylase 1** | 36663290.9 | 15625365.9 | 2.35 | 3.28E-03 | 2.22E-02 |
| **Calmodulin-like protein 3** | 12688.5703 | 4613.72618 | 2.75 | 3.28E-03 | 2.22E-02 |
| **Secretoglobin family 3A member 1** | 101785.721 | 15167.0479 | 6.71 | 3.28E-03 | 2.22E-02 |
| **C4b-binding protein alpha chain** | 48991.6786 | 108461.72 | 0.45 | 3.75E-03 | 2.39E-02 |
| **CD177 antigen** | 20675.2812 | 40383.9624 | 0.51 | 3.75E-03 | 2.39E-02 |
| **Peroxiredoxin-6** | 15935.584 | 9920.11174 | 1.61 | 3.75E-03 | 2.39E-02 |
| **Ras-related protein Rab-10** | 8322.0337 | 3541.32706 | 2.35 | 3.75E-03 | 2.39E-02 |
| **Arginase-1** | 76651.5558 | 133725.985 | 0.57 | 4.28E-03 | 2.48E-02 |
| **Neutrophil collagenase** | 440685.698 | 701174.871 | 0.63 | 4.28E-03 | 2.48E-02 |
| **Protein S100-A11** | 713525.122 | 615253.852 | 1.16 | 4.28E-03 | 2.48E-02 |
| **Choline transporter-like protein 4** | 126677.509 | 77769.1373 | 1.63 | 4.28E-03 | 2.48E-02 |
| **Cathepsin D** | 198963.266 | 113984.912 | 1.75 | 4.28E-03 | 2.48E-02 |
| **Kininogen-1** | 21013.9249 | 8984.9347 | 2.34 | 4.28E-03 | 2.48E-02 |
| **Histone H1.4** | 333376.043 | 59183.7839 | 5.63 | 4.28E-03 | 2.48E-02 |
| **Proteasome subunit alpha type-1** | 9919.49007 | 19786.786 | 0.50 | 4.88E-03 | 2.62E-02 |

| | | | | | |
|---|---|---|---|---|---|
| **Superoxide dismutase [Mn], mitochondrial** | 21067.7863 | 38149.753 | 0.55 | 4.88E-03 | 2.62E-02 |
| **Alpha-2-macroglobulin** | 2034071 | 3378909.52 | 0.60 | 4.88E-03 | 2.62E-02 |
| **Interleukin-1 receptor antagonist protein** | 330981.183 | 194570.063 | 1.70 | 4.88E-03 | 2.62E-02 |
| **Protein disulfide-isomerase A3** | 143947.871 | 83955.1973 | 1.71 | 4.88E-03 | 2.62E-02 |
| **Cysteine-rich secretory protein 3** | 246185.484 | 41431.2331 | 5.94 | 4.88E-03 | 2.62E-02 |
| **Nucleobindin-1** | 47066.9034 | 40777.2797 | 1.15 | 5.54E-03 | 2.87E-02 |
| **Immunoglobulin kappa variable 1-5** | 88145.4681 | 56517.0016 | 1.56 | 5.54E-03 | 2.87E-02 |
| **Serotransferrin** | 7085712.41 | 3150953.79 | 2.25 | 5.54E-03 | 2.87E-02 |
| **Annexin A3** | 1557305.71 | 2888687.15 | 0.54 | 6.28E-03 | 3.15E-02 |
| **Myosin regulatory light chain 12B** | 149424.214 | 178882.87 | 0.84 | 6.28E-03 | 3.15E-02 |
| **Glutathione S-transferase P** | 889380.977 | 579870.654 | 1.53 | 6.28E-03 | 3.15E-02 |
| **ADP-ribosyl cyclase/cyclic ADP-ribose hydrolase 2** | 52917.8508 | 88516.8513 | 0.60 | 7.10E-03 | 3.37E-02 |
| **Immunoglobulin kappa variable 4-1** | 306689.706 | 407058.563 | 0.75 | 7.10E-03 | 3.37E-02 |
| **Growth factor receptor-bound protein 2** | 10043.1027 | 5956.84579 | 1.69 | 7.10E-03 | 3.37E-02 |
| **Prosaposin** | 313739.145 | 165082.272 | 1.90 | 7.10E-03 | 3.37E-02 |
| **Putative elongation factor 1-alpha-like 3** | 89087.1724 | 31974.8342 | 2.79 | 7.10E-03 | 3.37E-02 |

| | | | | | |
|---|---|---|---|---|---|
| **Complement factor H** | 690888.001 | 1354862.13 | 0.51 | 8.01E-03 | 3.61E-02 |
| **Actin-related protein 2** | 78243.2463 | 118736.186 | 0.66 | 8.01E-03 | 3.61E-02 |
| **Peroxiredoxin-5, mitochondrial** | 301372.826 | 179253.727 | 1.68 | 8.01E-03 | 3.61E-02 |
| **Actin, alpha cardiac muscle 1** | 553254.233 | 303008.476 | 1.83 | 8.01E-03 | 3.61E-02 |
| **Mammaglobin-B** | 204535.012 | 69938.4011 | 2.92 | 8.01E-03 | 3.61E-02 |
| **Pulmonary surfactant-associated protein B** | 301608.08 | 194118.242 | 1.55 | 9.02E-03 | 3.98E-02 |
| **Lysozyme C** | 51205332.9 | 25704892.5 | 1.99 | 9.02E-03 | 3.98E-02 |
| **Bactericidal permeability-increasing protein** | 131438.534 | 205536.582 | 0.64 | 1.01E-02 | 4.29E-02 |
| **Alpha-1B-glycoprotein** | 234251.08 | 346696.012 | 0.68 | 1.01E-02 | 4.29E-02 |
| **Immunoglobulin heavy constant mu** | 3154705.14 | 1818917.92 | 1.73 | 1.01E-02 | 4.29E-02 |
| **Beta-2-glycoprotein 1** | 141766.125 | 36198.4924 | 3.92 | 1.01E-02 | 4.29E-02 |
| **BPI fold-containing family A member 2** | 546650.998 | 946788.556 | 0.58 | 1.13E-02 | 4.58E-02 |
| **BPI fold-containing family A member 1** | 977494.722 | 612461.195 | 1.60 | 1.13E-02 | 4.58E-02 |
| **Galectin-1** | 34849.2492 | 21222.9777 | 1.64 | 1.13E-02 | 4.58E-02 |
| **SH3 domain-binding glutamic acid-rich-like protein 3** | 64173.8647 | 37886.85 | 1.69 | 1.13E-02 | 4.58E-02 |
| **Albumin** | 112375225 | 57372623.4 | 1.96 | 1.13E-02 | 4.58E-02 |

| | | | | | |
|---|---|---|---|---|---|
| **Apolipoprotein A-II** | 24274.7787 | 14928.6992 | 1.63 | 1.27E-02 | 5.06E-02 |
| **Galectin-10** | 114448.957 | 60342.7501 | 1.90 | 1.27E-02 | 5.06E-02 |
| **Small proline-rich protein 3** | 80686.3061 | 42488.77 | 1.90 | 1.42E-02 | 5.60E-02 |
| **Transthyretin** | 1202.93956 | 4091.54353 | 0.29 | 1.45E-02 | 5.67E-02 |
| **Leukocyte elastase inhibitor** | 319999.457 | 570683.936 | 0.56 | 1.58E-02 | 5.92E-02 |
| **Myosin-9** | 880759.822 | 975444.605 | 0.90 | 1.58E-02 | 5.92E-02 |
| **Extracellular glycoprotein lacritin** | 300387.711 | 212536.421 | 1.41 | 1.58E-02 | 5.92E-02 |
| **Mucin-4** | 14906.677 | 8595.28534 | 1.73 | 1.58E-02 | 5.92E-02 |
| **Guanine nucleotide-binding protein G(I)/G(S)/G(T) subunit beta-2** | 16381.5848 | 6689.91251 | 2.45 | 1.58E-02 | 5.92E-02 |
| **IgGFc-binding protein** | 1772571.39 | 1067895.48 | 1.66 | 1.75E-02 | 6.45E-02 |
| **Protein-glutamine gamma-glutamyltransferase 2** | 14739.8473 | 7368.29918 | 2.00 | 1.75E-02 | 6.45E-02 |
| **Plastin-2** | 3880822.76 | 5007491.38 | 0.78 | 1.95E-02 | 6.85E-02 |
| **Superoxide dismutase [Cu-Zn]** | 35073.8334 | 38011.5141 | 0.92 | 1.95E-02 | 6.85E-02 |
| **Zinc-alpha-2-glycoprotein** | 5502428 | 4262512.34 | 1.29 | 1.95E-02 | 6.85E-02 |
| **Beta-microseminoprotein** | 302251.655 | 193705.482 | 1.56 | 1.95E-02 | 6.85E-02 |
| **Myeloid cell nuclear differentiation antigen** | 180085.969 | 102150.441 | 1.76 | 1.95E-02 | 6.85E-02 |

| | | | | | |
|---|---|---|---|---|---|
| **Parkinson disease protein 7** | 89720.2726 | 50768.4674 | 1.77 | 1.95E-02 | 6.85E-02 |
| **Phosphoglucomutase-2** | 10214.6896 | 17557.2413 | 0.58 | 2.16E-02 | 7.35E-02 |
| **Myeloperoxidase** | 11774628.2 | 18573029.5 | 0.63 | 2.16E-02 | 7.35E-02 |
| **Actin-related protein 2/3 complex subunit 2** | 165183.861 | 207296.553 | 0.80 | 2.16E-02 | 7.35E-02 |
| **Glutathione S-transferase A1** | 41813.6469 | 29878.9578 | 1.40 | 2.16E-02 | 7.35E-02 |
| **Histone H3.2** | 1181954.54 | 2023164.44 | 0.58 | 2.39E-02 | 7.78E-02 |
| **Fatty acid-binding protein, adipocyte** | 29363.6118 | 25076.1809 | 1.17 | 2.39E-02 | 7.78E-02 |
| **Protein S100-A7** | 51358.1588 | 31918.165 | 1.61 | 2.39E-02 | 7.78E-02 |
| **Nucleolin** | 14551.5637 | 8524.26289 | 1.71 | 2.39E-02 | 7.78E-02 |
| **Nucleoside diphosphate kinase B** | 12955.504 | 6437.31256 | 2.01 | 2.39E-02 | 7.78E-02 |
| **Serpin B3** | 3580.18653 | 462.200932 | 7.75 | 2.39E-02 | 7.78E-02 |
| **Carcinoembryonic antigen-related cell adhesion molecule 8** | 43751.3771 | 69723.9892 | 0.63 | 2.63E-02 | 8.20E-02 |
| **Beta-glucuronidase** | 17620.9833 | 22961.91 | 0.77 | 2.63E-02 | 8.20E-02 |
| **Cathepsin B** | 54983.6941 | 45540.3092 | 1.21 | 2.63E-02 | 8.20E-02 |
| **Leucine-rich alpha-2-glycoprotein** | 74776.564 | 55350.6389 | 1.35 | 2.63E-02 | 8.20E-02 |
| **NPC intracellular cholesterol transporter 2** | 298150.262 | 187338.496 | 1.59 | 2.63E-02 | 8.20E-02 |

| | | | | | |
|---|---|---|---|---|---|
| **Cofilin-1** | 1235773.97 | 687469.636 | 1.80 | 2.63E-02 | 8.20E-02 |
| **Hemopexin** | 1028417.61 | 859112.728 | 1.20 | 2.90E-02 | 8.92E-02 |
| **Mucin-5B** | 13905884 | 7961109.13 | 1.75 | 2.90E-02 | 8.92E-02 |
| **Phosphatidylethanolamine-binding protein 1** | 186200.408 | 142319.798 | 1.31 | 3.19E-02 | 9.61E-02 |
| **Apoptosis-associated speck-like protein containing a CARD** | 40664.5952 | 26466.8693 | 1.54 | 3.19E-02 | 9.61E-02 |
| **Heterogeneous nuclear ribonucleoprotein F** | 31731.3254 | 18260.8945 | 1.74 | 3.19E-02 | 9.61E-02 |
| **Purine nucleoside phosphorylase** | 315892.271 | 426844.38 | 0.74 | 3.51E-02 | 1.04E-01 |
| **Alpha-1-antichymotrypsin** | 301697.575 | 234324.319 | 1.29 | 3.51E-02 | 1.04E-01 |
| **Cystatin-D** | 51231.7472 | 9704.0512 | 5.28 | 3.51E-02 | 1.04E-01 |
| **Keratin, type I cytoskeletal 16** | 12549.7824 | 28161.9711 | 0.45 | 3.84E-02 | 1.10E-01 |
| **Proteasome subunit beta type-8** | 24805.6695 | 35634.2157 | 0.70 | 3.84E-02 | 1.10E-01 |
| **Cystatin-S** | 3417075.41 | 1707790.98 | 2.00 | 3.84E-02 | 1.10E-01 |
| **T-complex protein 1 subunit delta** | 544.916149 | 205.118545 | 2.66 | 3.84E-02 | 1.10E-01 |
| **Alpha-actinin-1** | 668354.427 | 893907.081 | 0.75 | 4.21E-02 | 1.17E-01 |
| **Immunoglobulin lambda variable 3-9** | 282165.092 | 212642.205 | 1.33 | 4.21E-02 | 1.17E-01 |
| **40S ribosomal protein SA** | 10833.8141 | 8147.33968 | 1.33 | 4.21E-02 | 1.17E-01 |

| | | | | | |
|---|---|---|---|---|---|
| **Complement C3** | 7158650.1 | 5215554.07 | 1.37 | 4.21E-02 | 1.17E-01 |
| **Peroxiredoxin-1** | 33712.6391 | 18847.9355 | 1.79 | 4.21E-02 | 1.17E-01 |
| **HLA class II histocompatibility antigen, DR alpha chain** | 4764.27763 | 2345.80963 | 2.03 | 4.21E-02 | 1.17E-01 |
| **Low affinity immunoglobulin gamma Fc region receptor III-A** | 86449.9658 | 150575.038 | 0.57 | 4.60E-02 | 1.24E-01 |
| **Transketolase** | 5408875.51 | 6869256.21 | 0.79 | 4.60E-02 | 1.24E-01 |
| **Integrin alpha-M** | 59998.654 | 38188.1877 | 1.57 | 4.60E-02 | 1.24E-01 |
| **Immunoglobulin lambda constant 2** | 449652.459 | 189365.504 | 2.37 | 4.60E-02 | 1.24E-01 |
